# Environmental Surface Monitoring as a Noninvasive Method for SARS-CoV-2 Surveillance in Community Settings: Lessons from a University Campus Study

**DOI:** 10.1101/2023.07.13.23292575

**Authors:** Sobur Ali, Eleonora Cella, Catherine Johnston, Michael Deichen, Taj Azarian

## Abstract

Environmental testing of high-touch objects is a potential noninvasive approach for monitoring population-level trends of SARS-CoV-2 and other respiratory viruses within a defined setting. We aimed to determine the association between SARS-CoV-2 contamination on high-touch environmental surfaces, community level case incidence, and university student health data. Environmental swabs were collected from January 2022 to November 2022 from high-touch objects and surfaces from five locations on a large university campus in Florida, USA. RT-qPCR was used to detect and quantify viral RNA, and a subset of positive samples was analyzed by viral genome sequencing to identify circulating lineages. During the study period, we detected SARS-CoV-2 viral RNA on 90.7% of 162 tested samples. Levels of environmental viral RNA correlated with trends in community-level activity and case reports from the student health center. A significant positive correlation was observed between the estimated viral gene copy number in environmental samples and the weekly confirmed cases at the university. Viral sequencing data from environmental samples identified lineages contemporaneously circulating in the local community and state based on genomic surveillance data. Further, we detected emerging variants in environmental samples prior to their identification by clinical genomic surveillance. Our results demonstrate the utility of viral monitoring on high-touch environmental surfaces for SARS-CoV-2 surveillance at a community level. In communities with delayed or limited testing facilities, immediate environmental surface testing may considerably inform epidemic dynamics.

**Graphical Abstract:** 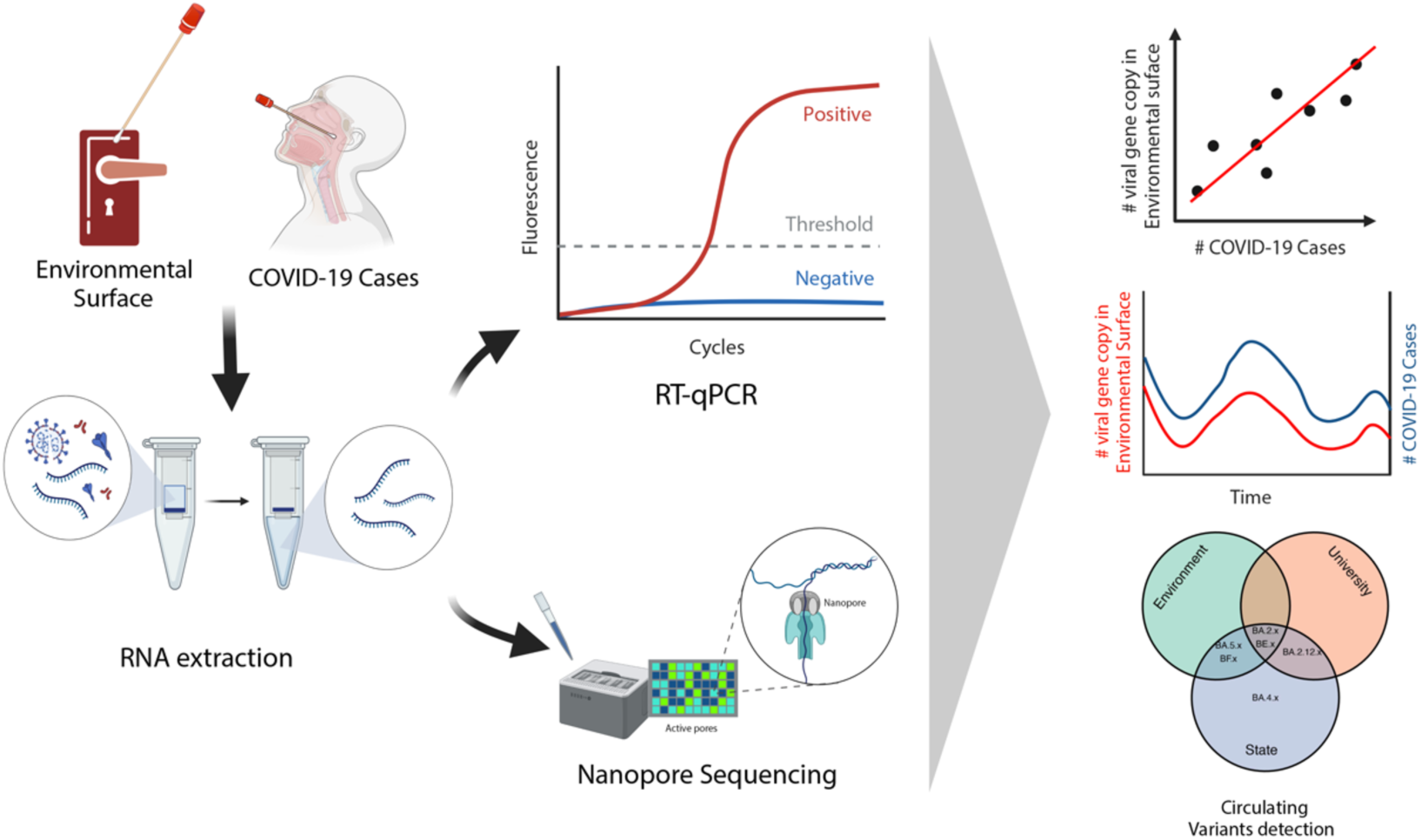

## Introduction

The rapid global spread of severe acute respiratory syndrome coronavirus 2 (SARS-CoV-2) represented a major public health emergency, and despite myriad control measures, community transmission is ongoing. Throughout the pandemic, monitoring SARS-CoV-2 activity has been a central component of the response. Monitoring activities have included testing frequency and positivity, morbidity and mortality tracking, viral genomic surveillance of emerging variants, and wastewater surveillance (Chen et al., 2022; Bowes et al., 2023). Despite substantial investment in these activities, surveillance “blind spots” were reported (Batista et al., 2022; Brito et al., 2022; Duma et al., 2022). Further, each approach requires evaluation of the utility, cost, equity, accessibility, representativeness, and timeliness. For example, while viral genomic surveillance provided the finest level of resolution in pandemic activity, it is costly and requires specialized equipment, which limited its accessibility in many settings.

Wastewater surveillance rose to popularity for its ability to provide a non-invasive, cost-effective, and efficient method for monitoring community level trends (Leifels et al., 2023). Wastewater surveillance detects the presence of SARS-CoV-2 RNA in wastewater before clinical cases are reported, allowing for early detection and response to potential outbreaks (Davó et al., 2021; Jahn et al., 2022; Karthikeyan et al., 2022; Wilhelm et al., 2022b). Similarly, environmental surveillance of high touch objects is non-invasive, less costly than clinical testing, and promises less biased sampling, especially for pathogens with a high proportion of asymptomatic or paucisymptomatic infections. Also, environmental surface monitoring reduces or eliminates issues related to participant recruitment, sample collection and operations logistics that can slow down or restrain clinical-testing programs (Arons et al., 2020; Lee et al., 2020). Targeted sampling of high-touch surfaces can be a potential pandemic surveillance strategy by identifying recent locations of currently infectious individuals (e.g., buildings or rooms), providing insight into infection load in a community setting and allowing monitoring of long-term trends.

The rapid evolution of the SARS-CoV-2 virus resulted in the emergence of variants of concern (VOCs) and variants of interest (VOIs) with increased infectivity, transmission, and immune evasion characteristics (Davies et al., 2021; Lucas et al., 2021). This underscored the necessity for ongoing monitoring of the emergence of novel variants. Thus, having reliable and timely detection of local prevalence of SARS-CoV-2 variants is crucial for developing effective public health measures. However, current strategies for variant detection, which mostly rely on virus genome sequencing of biospecimens obtained from clinical testing, face limitations such as high cost, inefficiency, and sampling bias due to systemic healthcare disparities, particularly in underserved communities (Lieberman-Cribbin et al., 2020; Reitsma et al., 2021; Brito et al., 2022). Compared to clinical genomic surveillance, environmental genomic surveillance for SARS-CoV-2 may enable the early detection of emerging variants of concern before they are identified through clinical genomic surveillance (Al Huraimel et al., 2020). Environmental genomic surveillance also provides a broader and more thorough overview of the prevalence and geographic distribution of SARS-CoV-2 in a population, capturing data from entire communities or geographical regions, which can assist in identifying viral hotspots, tracking trends over time, and guiding population-level decision-making (Ahmed et al., 2020).

Assessment of SARS-CoV-2 exposure risk is necessary for various non-healthcare social settings in universities that are potentially risky due to overcrowding. In such a setting, cross-contamination between employees, students, and high-touch surfaces can lead to indirect transmission via fomites (for Immunization, 2021; Heneghan et al., 2021; Kwon et al., 2023). By monitoring the presence of SARS-CoV-2 on surfaces in university settings, such as classrooms, libraries, and recreational areas, and understanding the dynamics of environmental contamination and transmission routes, targeted interventions can be implemented to prevent the spread of the virus within the university community. Effective environmental monitoring can complement other public health measures, such as testing and contact tracing, and contribute to a comprehensive approach to controlling the spread of SARS-CoV-2 in the university or similar setting such as healthcare facilities.

In this study, we conducted longitudinal environmental sampling from non-healthcare high-touch surfaces during the COVID-19 pandemic to measure SARS-CoV-2 contamination levels in different locations on the university campus. Our primary objectives were to evaluate the presence of viral RNA on environmental surfaces and the relationship between environmental surveillance results and clinically confirmed case data at the university level. Further, we carried out viral genome sequencing of these samples to investigate the feasibility of characterizing circulating lineages of SARS-CoV-2. Environmental public health surveillance studies such as this have the potential to provide insight into infection load and trends in preparation for future outbreaks.

## Materials and methods

### Environmental surface swab samples collection

We collected environmental surface swabs from high-touch areas in non-healthcare settings including, door handles from five locations: the Student Union (SU), Library, Gymnasium, Physical Sciences Building, and Business Administration Buildings of a large university in central Florida with 35,000-40,000 students. We collected samples weekly or biweekly from January 27, 2022, to November 20, 2022. Sterile cotton swabs (Fisher Scientific, USA) were moistened in viral transport media DNA/RNA Shield™ (Zymo Research Corp), then swabbed the outside and inside surface of the door handle horizontally or vertically, rotating the swab throughout. The swabs were placed in a sterile tube with three mL viral transport media and stored in sample carrier box with icebag during the sampling. Following the collection, the swabbed surfaces were wiped with 70% ethanol for sterilization. The samples were transported to the lab and stored at -80 °C until further processing. Hand hygiene, face mask and glove changes were performed during every sample collection.

### Positive and negative control

Several controls were used throughout the process to prevent false positive results and cross-contamination: 1) Sample site negative control: Fresh cotton swab placed in DNA/RNA shield at the sampling site, 2) RNA extraction negative control: Fresh cotton in fresh DNA/RNA shield at the lab during extraction, 3) Elution negative control: Only the elution buffer through the spin column, 4) qPCR non-template negative control: Nuclease-free water as a negative control during qPCR. 5) qPCR Positive Control: SARS-CoV-2 N1 gene fragment for PCR positive control in each run.

### RNA extraction and RT-qPCR

Viral RNA extraction was performed from 200 µl samples using QIAmp Viral RNA mini kit (Qiagen) and eluded in 40 µl elution buffer. RT-qPCR was performed to detect the SARS-CoV-2 RNA using primer/probe sets recommended by the US CDC that target a region of the nucleocapsid (N1) gene (Lu et al., 2020). RT-qPCR reactions were conducted in a 20 μl reaction using 5 µl 4X TaqPath master mix (Thermo Fisher Scientific, Massachusetts, USA), 0.5 uM each of 2019-nCoV_N1 (CDC) qPCR probe (5’-FAM-ACCCCGCATTACGTTTGGTGGACC-BHQ1-3’), forward primer (5’-GACCCCAAAATCAGCGAAAT-3’), and reverse primer (5’-TCTGGTTACTGCCAGTTGAATCTG-3’), 8.5μL of molecular-grade H_2_O, and 5 μl of template RNA. RT-qPCR was performed on a CFX Opus 96 instrument (Bio-Rad Laboratories, Hercules, California, USA) with the following conditions: initial incubation at 25°C for 2 min; reverse transcription step at 50°C for 15 min, followed by polymerase activation at 95°C for 2 min, and finally, 35 cycles of amplification at 95°C for 15s and 55°C for 30s. All samples were run in triplicate, including positive and negative controls. A sample was considered positive if at least one of the triplicates was amplified with a Ct below 35 in the N1 gene assay. Quantification of viral RNA in the environmental samples was based on a standard curve generated from serial dilutions of Synthetic SARS-CoV-2 RNA (ATCC VR-3276SD, Manassas, VA) with known concentration and converted to genome copy number per square centimeter (gc/cm^2^) of the surface genomic units per ten square centimeters of surface.

### SARS-CoV-2 genome sequencing from environmental surface

SARS-CoV-2 genome sequencing was conducted following the modifications to the ARTIC Network Protocol (v2) (Coil et al., 2021) to optimize sequencing of environmental samples. In brief, we conducted random hexamer primed reverse transcription and amplified cDNA using ARTIC v4.1 primers, which tile the entire viral genome with overlapping 400 bp fragments. We concentrated the amplified PCR products using the AMPure™ XP (Agencourt, Beckmann-Coulter, USA) magnetic bead. The sequence library was prepared using the Oxford Nanopore Native Barcoding kit (EXP-NBD104) and Ligation Sequencing Kit (SQK-LSK109). Libraries were run on ONT R9.4.1 flow cells on the GridION sequencing platform. Viral genome assembly was performed in two steps (using default parameters) following the ARTIC Network bioinformatics protocol (https://artic.network/ncov-2019/ncov2019-bioinformatics-sop.html). The gupplyplex script was used for quality control and filtering of reads 400 bp followed by assembly with medaka using Wuhan-Hu-1 reference genome (GenBank accession number MN908947.3). From the sequencing data circulating SARS-CoV-2 variants were identified using Freyja (version 1.3.11) (Karthikeyan et al., 2022).

### Clinical sample processing and sequencing of SARS-CoV-2 genome

Swab samples were collected from individuals seeking care at university health center during the study period. Collected swabs were placed in Zymo Research DNA/RNA shield and stored at 4°C until RNA extraction. RNA extraction for all samples was preformed using the QIAamp 96 virus QIAcube HT kit automated platform. RT-qPCR was performed following using CDC recommended protocol targeting N1 gene segment. SARS-CoV-2 viral genome sequencing and analysis was performed following the protocol as described previously (Hassouneh et al., 2023). The weekly distribution of variants was plotted over time.

### COVID-19 confirmed case and viral genome sequencing data

University COVID-19 weekly case data from January 2022 to November 2022 were collected from the student health center. Florida and Orange County 7-days moving averages cases data were collected from The New York Times. (2021). Coronavirus (Covid-19) Data in the United States. Retrieved [February 2023], from https://github.com/nytimes/covid-19-data. SARS-CoV-2 genomes generated from Florida state from January 2022 to November 2022 were downloaded from GISAID (www.gisaid.org, accessed on 15 June 2023 **Supplementary Table 1**). Assigned lineages metadata of these genomes were used to determine the weekly distribution of identified variants.

### Statistical analysis

All statistical analyses were performed in RStudio version 2023.3.0.386 (Posit Team, 2023) and *p-value* < 0.05 is considered statistically significant. The slope and intercept case numbers and RNA copy numbers were calculated using linear regression. The strength of a linear association was assessed using Spearman’s non-parametric correlation coefficient test due to non-normal distribution *p* < 0.05 with Shapiro-Wilk test. A Cross-correlation test was performed to examine the time-lagged association between weekly clinical COVID-19 cases in the university and environmental surface SARS-CoV-2 viral RNA gene copy. The causality was examined using the Granger causality test using the *grangertest* function in R, and significance was indicated when *P* < 0.05. All plots were generated with “ggplot” package (Wickham et al., 2016) in RStudio.

## Results

### Detection of viral RNA on environmental surface

We tested 162 swab samples from five locations between January 2022 and November 2022. Upon analyzing these samples, we found that 90.7% (147/162) of environmental surfaces tested positive for the N1 gene of SARS-CoV-2 based on results of RT-qPCR. Among the tested locations, a higher positivity rate was found in the Student Union and Library (Figure 1). The RT-qPCR Ct values of the positive samples ranged from 26.8 to 34.9. To better understand the viral gene copy on the environmental surfaces, we used a standard curve generated by synthetic SARS-CoV-2 genome to convert the Ct values into gene copy per square centimeter (gc/cm^2^), finding that viral gene copy ranged from 0.4 to 60.6 gc/cm^2^ (Table 1). Additionally, we observed similar patterns in the log value of gene copy numbers among sampling sites, except for the physical science building, where the viral gene copy was relatively lower than the other four sites (Figure 2).

**Figure 1:**
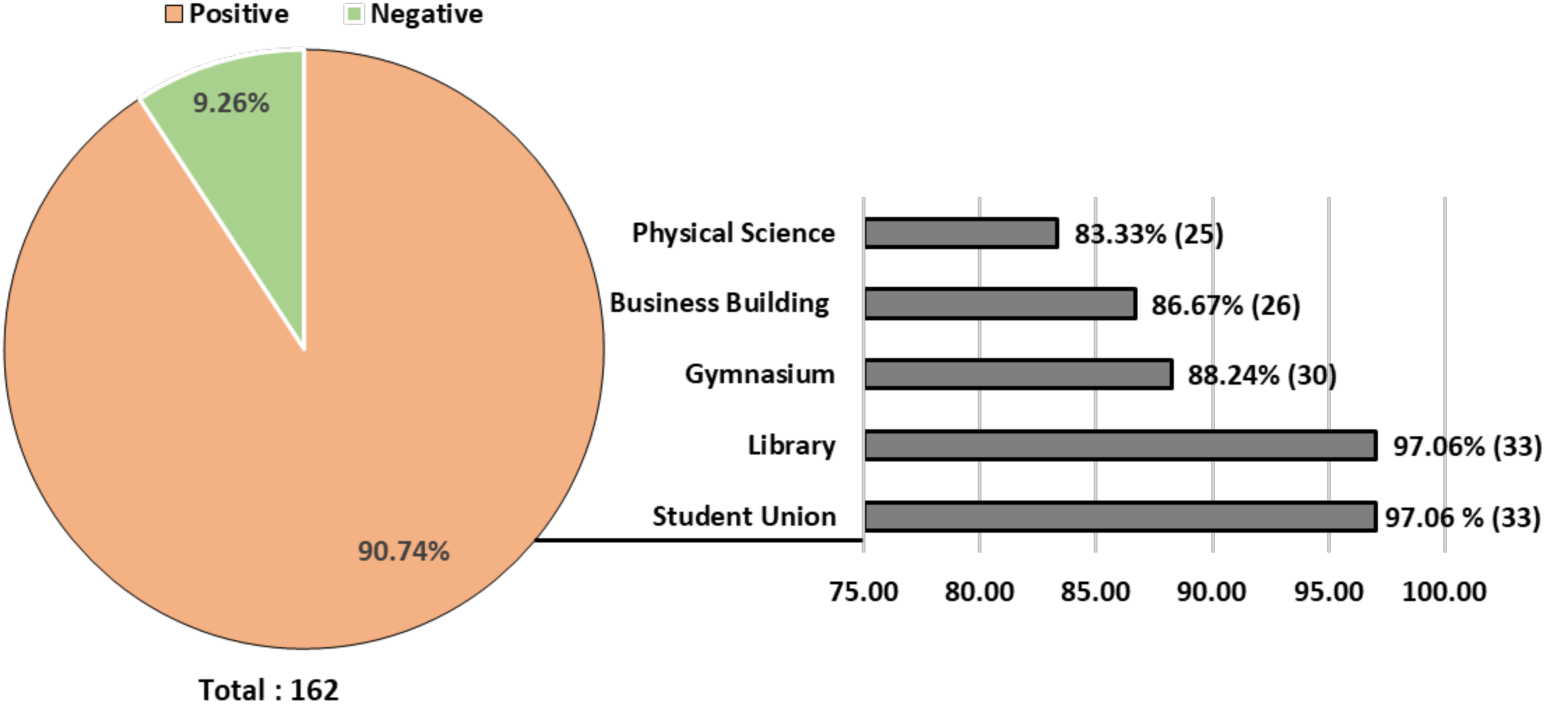
Distribution of positive samples for SARS-CoV-2 RNA in 162 samples collected at the University of Central Florida, Orlando, from five different locations. Of the 162 samples, 147 were positive for viral genome detection. Raw numbers and precents are denoted.

**Figure 2:**
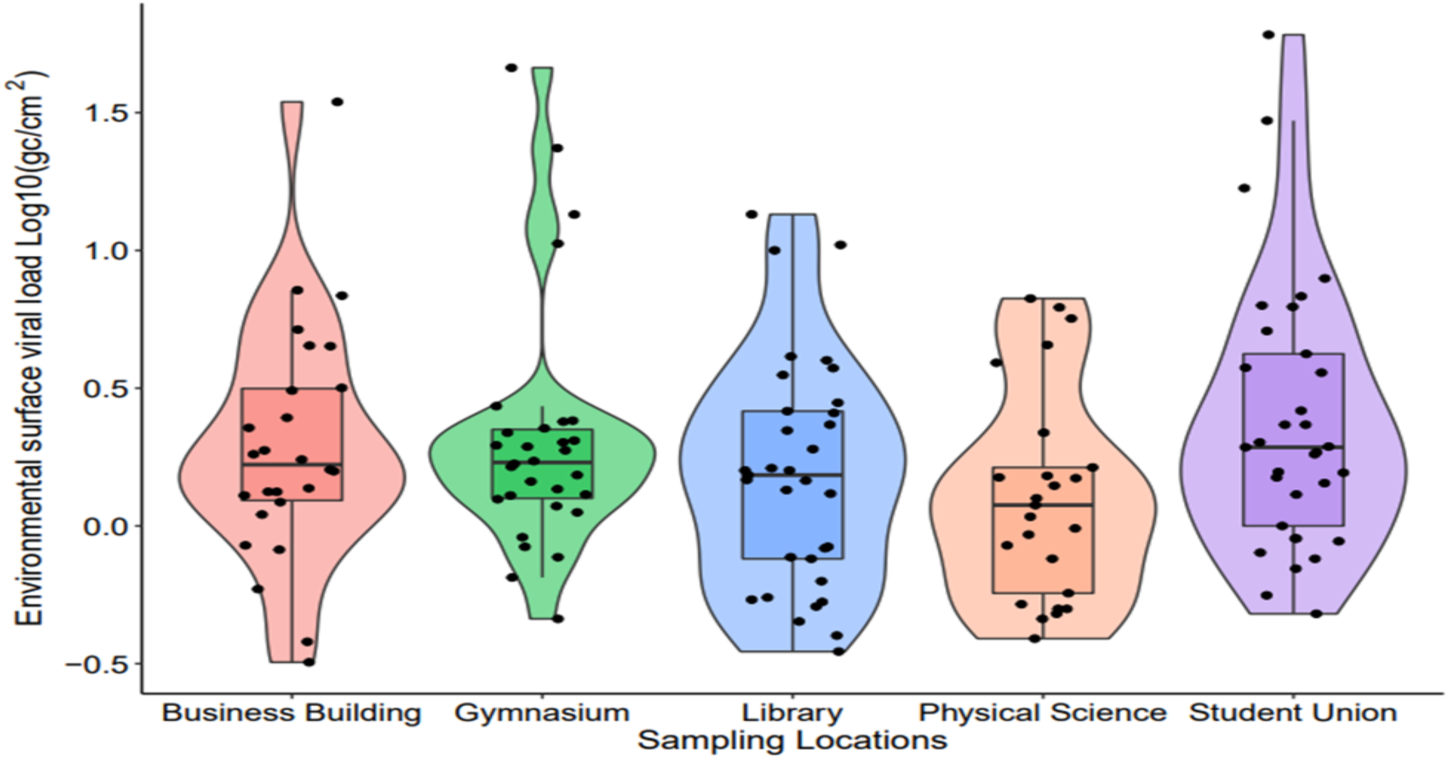
The environmental surface viral RNA concentration in Log10 of genome copy (N1 gene) number per square centimeter (gc/cm2) (Y-axis) at different sampling locations (X-axis) on the University campus represented by violin plots, and box plots. The color of each violin plot and box plot indicates the corresponding sampling location. The violin plots show the data distribution, with wider areas representing higher frequency. The box plots show the median and quartiles of the data. Note, locations were intentionally omitted from the x-axis.

**Table 1:** Viral Gene Copy number per square (GC/cm2) centimeter in five locations of the University of Central Florida campus during the study period.

| **Date of Sample Collection** | **Student Union (GC/cm<sup>2</sup>)** | **Library (GC/cm<sup>2</sup>)** | **Gymnasium (GC/cm<sup>2</sup>)** | **Business Building (GC/cm<sup>2</sup>)** | **Physical Science (GC/cm<sup>2</sup>)** |
| --- | --- | --- | --- | --- | --- |
| 1/27/2022 | Absent | Absent | 2.18 | Not Tested | Not Tested |
| 2/3/2022 | 4.21 | 2.22 | 2.39 | Not Tested | Not Tested |
| 2/9/2022 | 6.31 | 2.57 | 1.12 | Not Tested | Not Tested |
| 2/18/2022 | 1.93 | 0.55 | 0.65 | Not Tested | Not Tested |
| 2/25/2022 | 0.48 | 0.77 | Absent | Absent | 1.63 |
| 3/4/2022 | 1.43 | 0.40 | Absent | Absent | 0.76 |
| 3/18/2022 | 1.00 | 0.51 | 1.25 | 1.74 | 1.49 |
| 3/24/2022 | 0.88 | 0.35 | 0.46 | 0.85 | 0.93 |
| 3/31/2022 | 2.33 | 0.45 | Absent | 0.32 | Absent |
| 4/8/2022 | 1.94 | 0.84 | 2.04 | 1.33 | 0.57 |
| 4/15/2022 | 1.57 | 3.74 | 1.18 | 4.51 | 1.40 |
| 4/22/2022 | 2.33 | 1.35 | 0.84 | 0.59 | 0.52 |
| 5/6/2022 | 1.82 | 0.83 | 1.45 | Absent | Absent |
| 5/13/2022 | 3.60 | 1.46 | 23.51 | Absent | Absent |
| 5/20/2022 | 3.75 | 1.90 | 10.58 | 3.10 | 1.08 |
| 6/3/2022 | 6.81 | 1.59 | 2.26 | 2.27 | 2.18 |
| 6/10/2022 | 5.10 | 2.80 | 1.88 | 1.88 | 0.98 |
| 6/22/2022 | 16.82 | 4.12 | 1.64 | 1.22 | 1.52 |
| 7/1/2022 | 60.59 | 10.47 | 2.72 | 6.85 | 1.26 |
| 7/8/2022 | 29.57 | 3.99 | 0.77 | 2.47 | 5.66 |
| 7/15/2022 | 6.24 | 13.51 | 46.00 | 4.49 | 3.91 |
| 7/22/2022 | 7.91 | 10.00 | 13.50 | 1.29 | Absent |
| 8/8/2022 | 1.30 | 1.59 | Absent | 1.60 | 0.50 |
| 8/18/2022 | 0.70 | 2.33 | 1.30 | 1.37 | 1.19 |
| 8/25/2022 | 1.85 | 2.61 | 1.68 | 1.58 | 0.39 |
| 9/8/2022 | 1.56 | 3.53 | 1.96 | 7.17 | 4.54 |
| 9/16/2022 | 1.50 | 1.53 | 1.53 | 1.33 | 0.46 |
| 9/26/2022 | 0.80 | 1.47 | 1.72 | 3.17 | 0.85 |
| 10/7/2022 | 2.01 | 1.31 | 1.94 | 0.38 | 1.50 |
| 10/14/2022 | 0.56 | 0.54 | 1.29 | 34.57 | 0.48 |
| 10/24/2022 | 0.90 | 0.63 | 0.91 | 5.16 | 6.21 |
| 10/31/2022 | 0.76 | 0.53 | 2.01 | 1.82 | 6.69 |
| 11/16/2022 | 2.62 | 1.62 | 1.36 | 0.82 | 0.50 |
| 11/23/2022 | 0.90 | 0.76 | 2.41 | 1.10 | Absent |

### Trend of longitudinal viral gene copy on environmental surface and COVID-19 cases

The study was initiated at the end of January 2022 when COVID-19 cases associated with the Delta lineages were declining, and the Orange County region reported around 3,000 infections, while Florida had reported over 30,000 cases. By the end of February, cases declined; however, a subsequent increase in cases due to the Omicron lineage was reported between May and September 2022. Case trends in Florida and Orange County followed similar pattern (Figure 3A). During the early period of the study, the university had approximately 60 weekly confirmed cases. By the end of February, cases identified by the university student health center declined to below 10 cases weekly. An increase in cases was observed during the Omicron variant wave, between May and September. Throughout the study period, the confirmed cases in the university followed a similar trend to the cases reported in Florida state and Orange County (Figure 3B). The environmental surface swab results showed that in the early months of January and February, there was a higher viral gene copy on the surfaces, which declined as the number of confirmed cases decreased on the campus. However, during the Omicron wave, viral gene copy was sharply increased on the environmental surfaces (Figure 3C). Overall, the viral gene copy on the environmental surfaces exhibited a similar pattern with the confirmed cases on campus.

**Figure 3:**
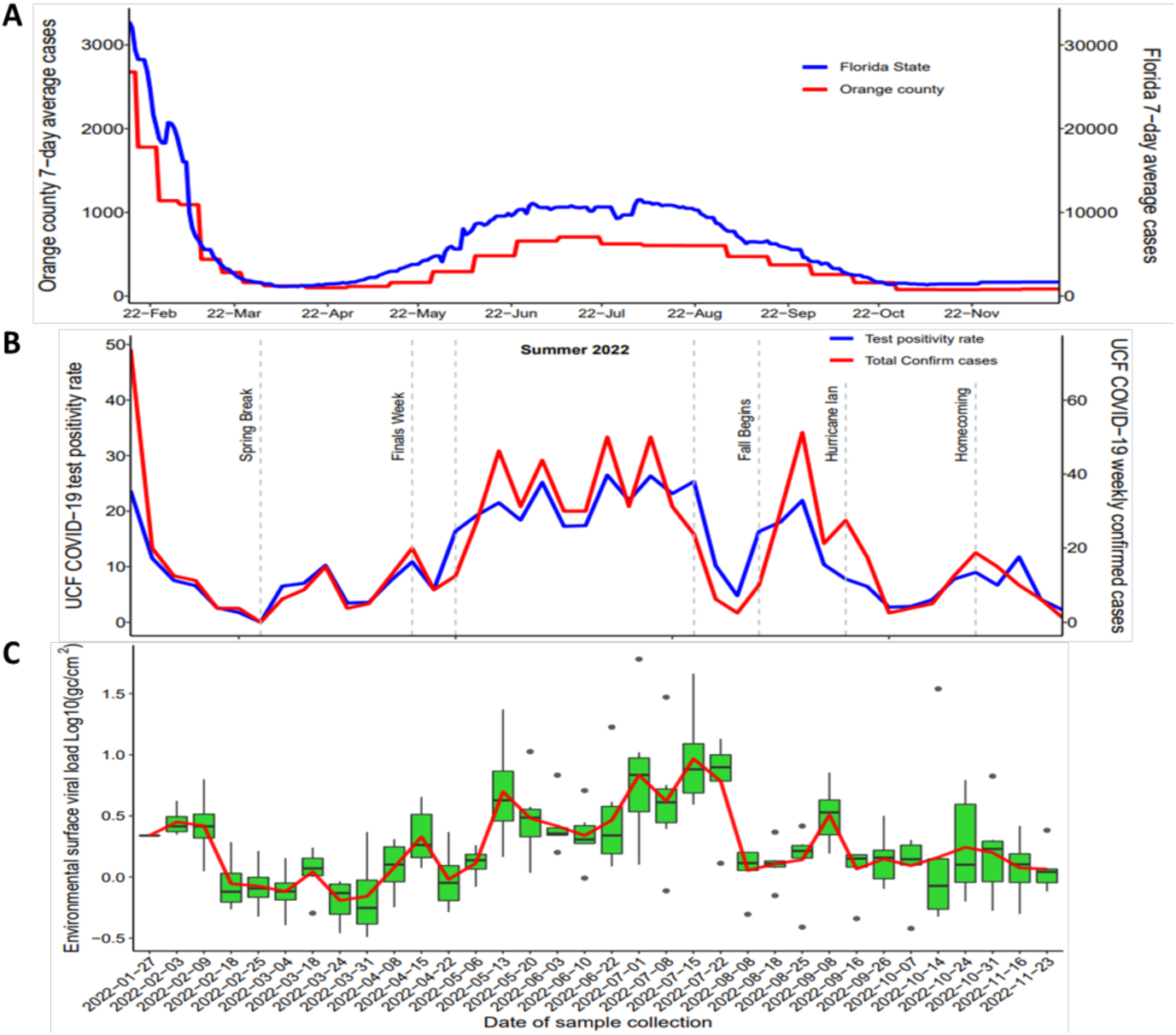
Comparison of the trend of COVID-19 confirmed cases and environmental surface viral gene copy. (A) Confirmed cases in the 7-day moving average of SARS-CoV-2 infection from 27 January 2022 to 27 November 2022 in Florida are shown in the y-axis on the right (red line) and in Orange shown in the y-axis on the left (blue line). (B) The trends of COVID-19 weekly confirmed cases and test positivity rate from 27 January 2022 to 23 October 2022. The x-axis represents the time, the y-axis on the left (blue line) represents the COVID-19 weekly test positivity rate, and the y-axis on the right (red line) represents the COVID-19 weekly confirmed cases. Dashed lines with text annotations indicate notable events during the study period, which may have influenced the observed trends. (C) Viral gene copy trend in environmental surfaces. Boxplots show the weekly aggregated viral gene copy distribution in log10 of genome copy (N1 gene) numbers per square centimeter (gc/cm2) across the five different sample collection sites. The red line shows the average viral gene copy trend over time.

### Association between viral genome copy number and confirmed cases

We used the Spearman correlation coefficient to assess the correlation between COVID-19 cases and viral gene copy on campus environmental surfaces. The Shapiro-Wilk normality test revealed that both variables did not meet the assumption of normality, and so a non-parametric test was employed instead. The Spearman correlation coefficient is a robust measure of association that does not assume a linear relationship or normal distribution of the data. Significant positive correlations between SARS-CoV-2 gene copy number on environmental surface and confirmed COVID-19 cases in the university (Spearman, p < 0.05), indicating that an increase in COVID-19 cases was associated with an increase on viral gene copy in the environmental surface (Figure 4). The Spearman correlation coefficient (rho) value was 0.629, indicating a moderate positive correlation between the two variables. The results of the Granger causality test showed that the test statistic (F) was 5.7105 with a p-value (Pr(>F)) of 0.023, indicating a statistically significant relationship between the log-transformed viral gene copy in environmental surfaces and weekly confirmed COVID-19 cases at the 0.05 significance level. We also examined the cross-correlation between viral gene copy and weekly case data to determine the lag between the two variables. We found that the highest correlation between weekly COVID-19 cases and viral gene copy occurs at a slight lag (0.02 weeks) (Supplementary Fig S1).

**Figure 4:**
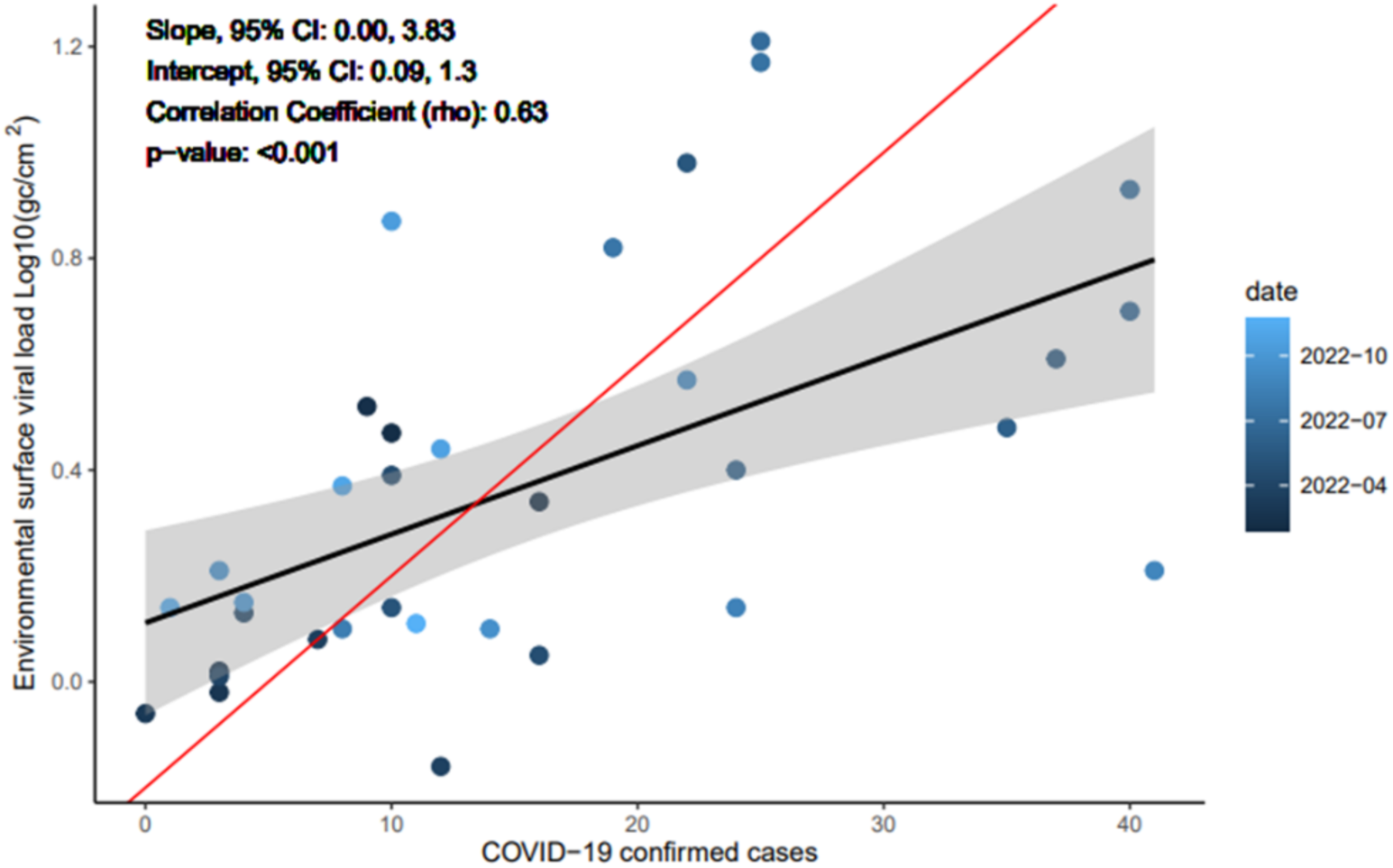
Relationship between environmental surface viral gene copy and COVID-19 confirmed cases. The plot displays the relationship between the average environmental surface viral gene copy measured in log10 of gene copy (N1 gene) number per square centimeter (gc/cm2) (Y-axis) and the number of weekly confirmed COVID-19 cases across different dates (X-axis). Each data point represents a particular week’s average viral gene copy and weekly COVID-19 case count on the university campus. Data points are colored by the date of sample collection, ranging from dark blue (earliest date) to light (latest date). The black line represents the linear regression line fit to the data points, with the 95% confidence interval shaded in gray. The slope and intercept of the regression line, its 95% confidence interval, Spearman’s rank correlation coefficient (rho), and the p-value are denoted. The red line represents perfect agreement (unity lines).

**Supplementary Figure 1:**
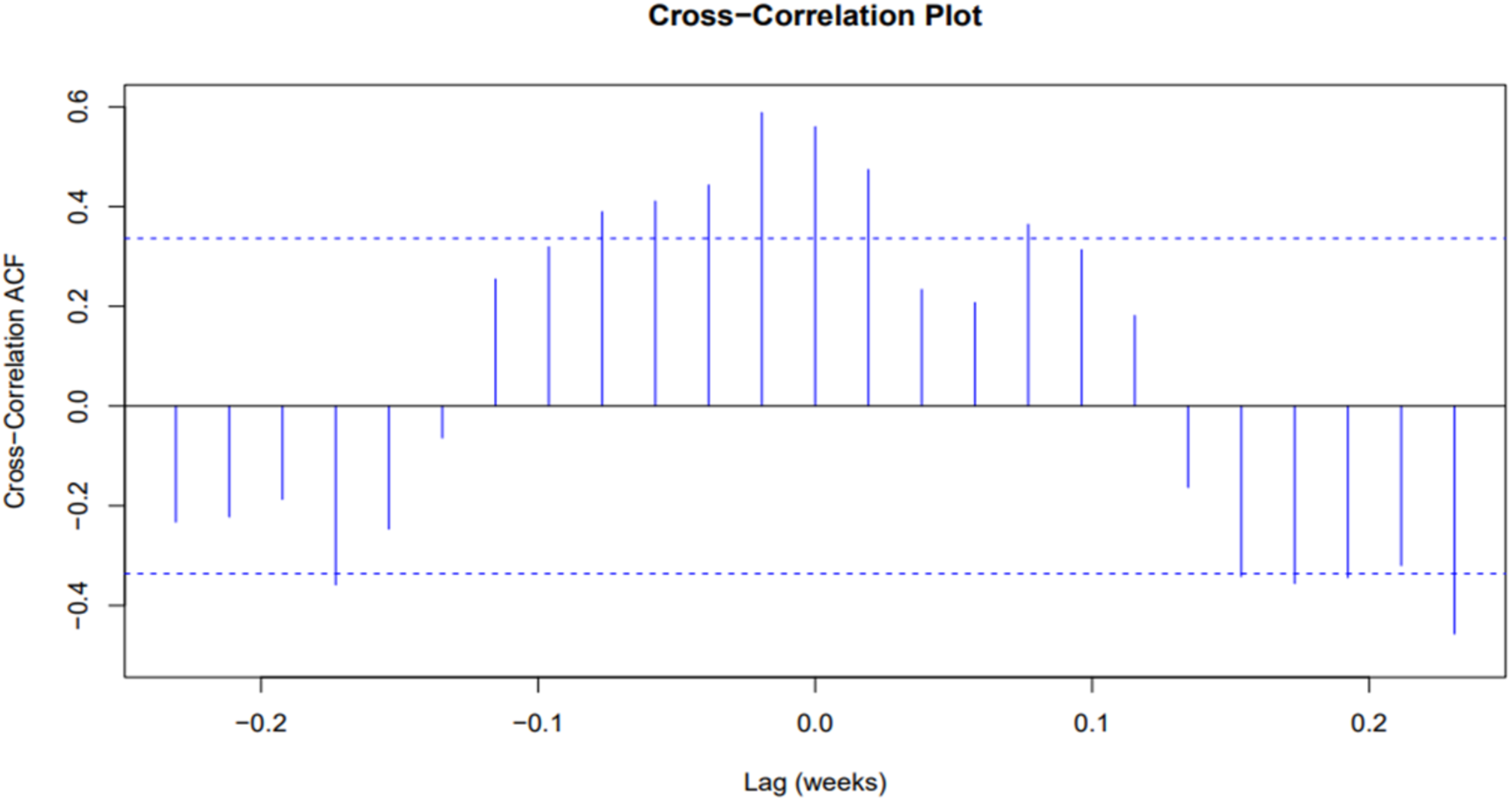
Cross-Correlation Function between Weekly COVID-19 Cases and Viral gene copy. The x-axis represents the lag in weeks, while the y-axis represents the correlation, measuring the strength and direction of the linear relationship. The solid line at y = 0 serves as a reference, representing no correlation between the two variables. The height of the solid line at each lag indicates the strength of the correlation.

### Detection of SARS-CoV-2 variants in environmental surface

We successfully sequenced 465 SARS-CoV-2 genomes from the university health center samples. In addition, 73,446 viral genome sequences from Florida cases were downloaded from GISAID. The distribution of variants over time was co-visualized, demonstrating temporal concordance in the composition of variants observed at the state and university level (Figure 5). To investigate the feasibility of variant detection from environmental samples, we carried out SARS-CoV-2 amplicon-based viral metagenomic sequencing from 15 environmental surface samples distributed over the study period (Sup Table1). Environmental sample sequencing coverage was negatively associated with Ct values, as lower Ct value samples yielded higher coverage (R = -0.9, p-value < 0.001, Supplementary Fig. S2). Variant composition of environmental viral sequencing data was assigned using Freyja and results were compared to contemporaneously circulating lineages identified in university and state level data. The result showed high concordance in detected SARS-CoV-2 variants between environmental surfaces and viral sequencing data from positive cases at the university health center and state (Table 2, Figure 5). In five instances, environmental viral genomic monitoring identified variants that were found in state-level data but not among university health center samples. In March 2022, we identified the BA.5.x lineage in sequencing data from two surface samples, despite not being detected in clinical genomic surveillance at the university and state levels.

**Table 2:** Distribution of SARS-CoV-2 lineages during the study period in university environmental samples, university health center clinical samples and Florida state. Bolded variants are those that were concordantly identified in viral genomic surveillance data from all three sites.

| Weeks | Identified Variants |  |  |
| --- | --- | --- | --- |
|  | Environmental Swab <sup>a</sup> (%) | University (%) | Florida State <sup>b</sup> (%) |
| 2022-W09 <sup>c</sup> | B.1.617 (14.7), BA.5.x (65.13) | BA.1.x (100) | BA.1.x (81.8), BA.2.x (17.4) |
| 2022-W11 | BA.5.x (99.6) | BA.1.x (25), BA.2.x (75) | BA.1.x (46.2), BA.2.x (52.3) |
| 2022-W20 <sup>c</sup> | **BA.2.x (90.8)**, BA.5.x (3.7) | BA.2.12.x (50), **BA.2.x (41.7)**, BG.x (8.3) | **BA.2.x (35.1)**, BA.2.12.x (59.9), BA.5.x (2.5) |
| 2022-W25 <sup>c</sup> | **BA.2.12.x (67.0)**, **BA.2.x (4.2)**, **BA.5.x (11.6)**, BG.x (9.5) | **BA.2.12.x (25)**, BA.4.x (12.5), **BA.5.x (50)**, BE.x (12.5) | BA.2.x (7.5), **BA.2.12.x (40.2)**, BA.4.x (13.2), **BA.5.x (32.3)**, BE.x (2.6), BF.x (3.5) |
| 2022-W26 <sup>c</sup> | **BA.2.12.x (1.4)**, BA.2.x (1.6), **BA.4.x (1.1)**, **BA.5.x (79.2)**, XAS (7.6) | **BA.2.x (4.3)**, **BA.2.12.x (34.8)**, **BA.4.x (13)**, **BA.5.x (34.8)**, **BE.x (8.7)**, BF.x (4.3) | **BA.2.x (4.2)**, **BA.2.12.x (28.1)**, **BA.4.x (12.8)**, **BA.5.x (42.8)**, **BE.x (6.7)**, BF.x (4.3) |
| 2022-W28 | **BF.x (39.2)**, **BA.5.x (56.1)** | BA.2.12.x (16), BA.4.x (24), **BA.5.x (48)**, BE.x (8), **BF.x (4)** | BA.2.12.x (10), BA.4.x (16.5), **BA.5.x (57.6)**, BE.x (6.5), **BF.x (7.1)** |
| 2022-W29 | BA.2.x (19.1), **BA.4.x (32.6)**, **BA.5.x (44.7)** | BA.2.12.x (5), **BA.4.x (15)**, **BA.5.x (60)**, **BE.x (5)**, BF.x (15) | BA.2.12.x (6), **BA.4.x (14)**, **BA.5.x (66.4)**, **BE.x (6.1)**, BF.x (6.1) |
| 2022-W38 <sup>c</sup> | BF.7 (93.3), **BA.5.x (5.6)** | BA.4.x (33.3), **BA.5.x (50)**, BE.x (8.3), BQ.1.x (8.3) | BA.4.x (12.6), **BA.5.x (65.4)**, BE.x (6.1), BF.x (13) |
| 2022-W39 | **BA.5.x (98.9)** | **BA.5.x (100)** | BA.4.x (13.4), **BA.5.x (61.4)**, BQ.1.x (3.1), BE.x (2.9), BF.x (14.1) |
| 2022-W40 | **BA.5.x (76.8)**, **BF.x (22.2)** | **BA.5.x (33.3)**, **BF.x (33.3)**, BQ.1.x (33.3) | **BA.4.x (10.6)**, **BA.5.x (60.2)**, BQ.1.x (7), BF.x (16.2) |
| 2022-W41 <sup>c</sup> | BA.5.x (96.7), BE.x (2) | BA.4.x (100) | BA.4.x (8.9), BA.5.x (59.2), BQ.1.x (10.5), BE.x (2.9), BF.x (14.5) |

Notes: <sup>a</sup> Identified lineages in environmental swab samples with their abundance proportion from the university on the designated weeks. Here lineages with abundance up to 1% is listed and details results were provided in Supplementary Table 2.
<sup>b</sup> Identified lineages in Florida State from available sequence data from GISAID in designated weeks. We listed all the lineages here with a proportion >2.5%.
<sup>c</sup> Instances where viral genomic monitoring identified variants that were found in state-level data but not among university health center samples

**Figure 5:**
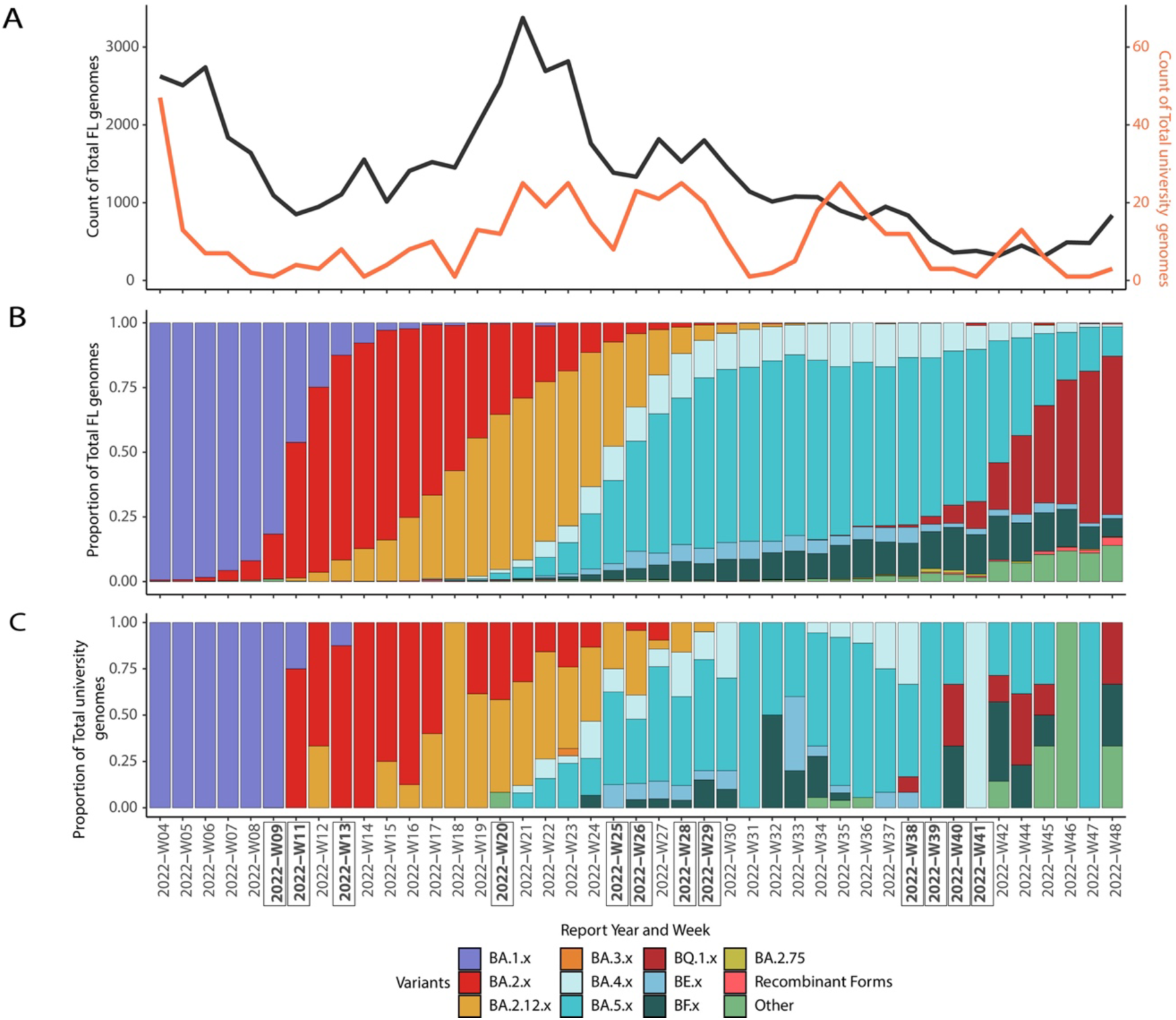
Lineage distribution identified by SARS-CoV-2 genome sequencing. (A) Total number of sequences analyzed from Florida state and university campus during the study period. (B) Distribution of lineages (y-axes) over time (x-axis) in proportion of sequenced isolates stratified by variant in Florida state and (C) university campus. Remaining lineages are grouped into “Other”. The weeks of environmental swab sample sequencing is annotated as bold.

**Supplementary Fig. S2:**
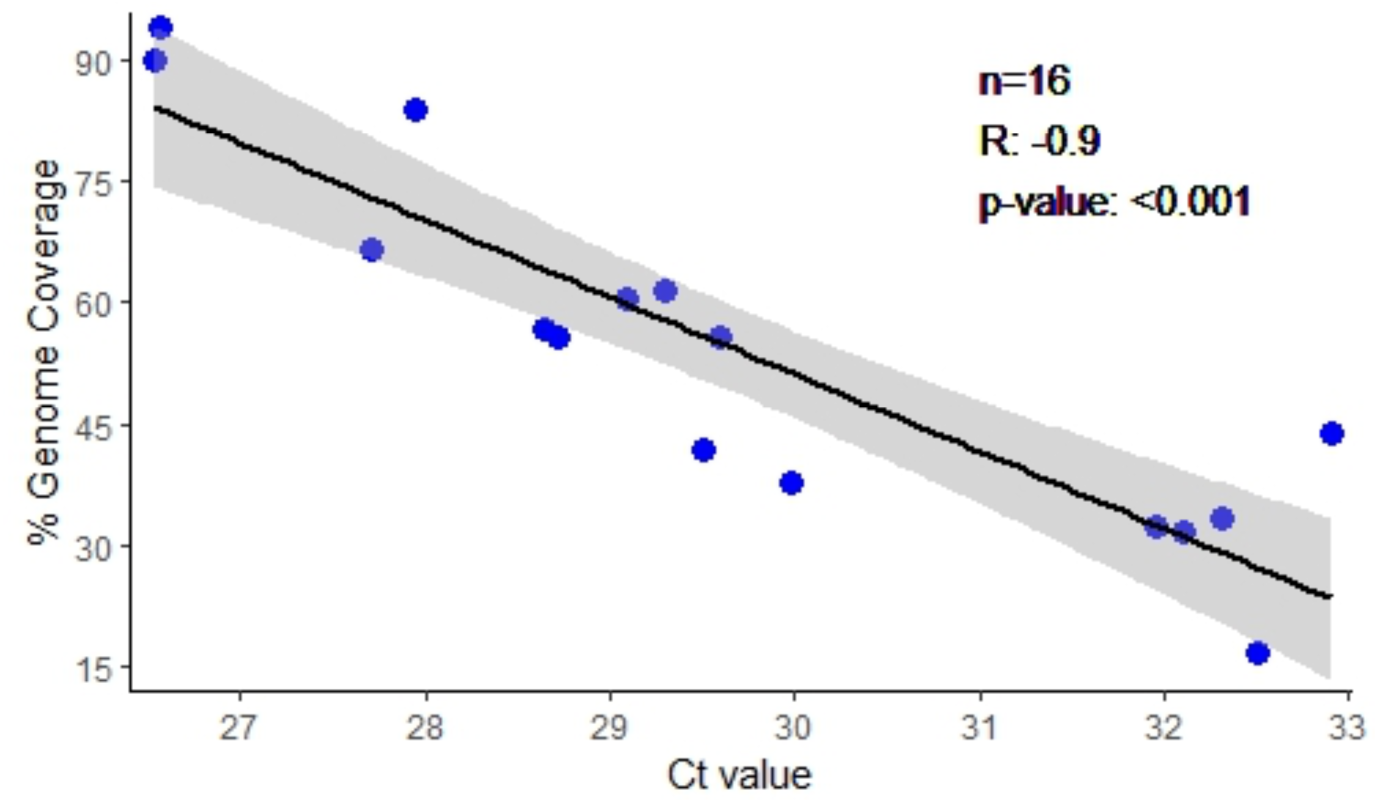
Correlation between SARS-CoV-2 viral genome coverage and the Ct value of 16 environmental samples used for viral metagenomic sequencing.

**Supplementary Table 1:**
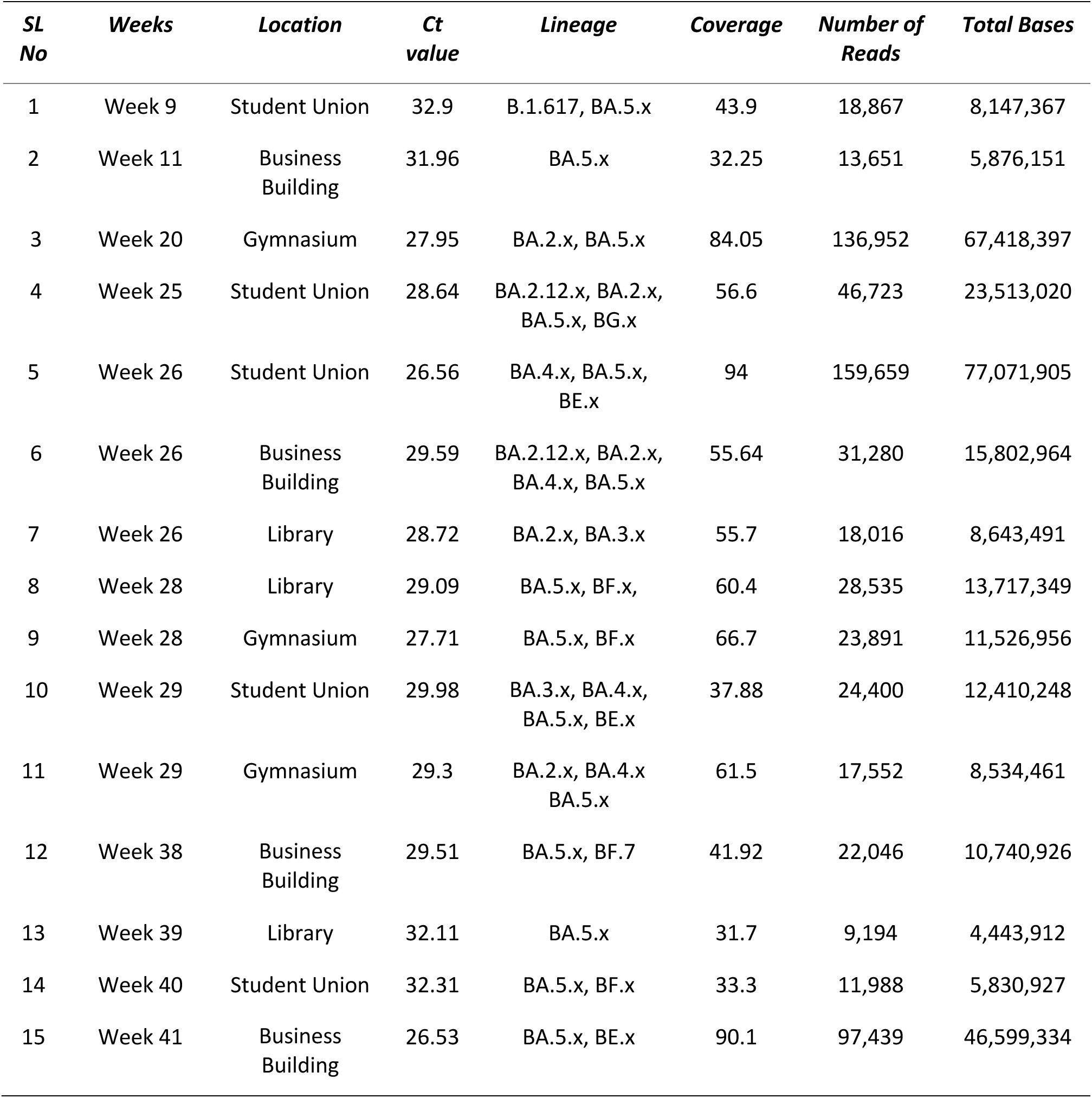
Environmental surface SARS-CoV-2 genome sequence results summary.

## Discussion

Environmental surveillance, in conjunction with individual clinical testing, is increasingly being applied by public health agencies to detect outbreaks, monitor pathogen evolution, and identify community-level trends (Anon, n.d.; Jahn et al., 2022; Smyth et al., 2022). More broadly, environmental monitoring may assist public health in making data-driven decisions about COVID-19 intervention and resource allocation. We aimed to detect SARS-CoV-2 in non-clinical high-touch environmental surfaces in a university setting. Our findings show that employing environmental surface monitoring in a defined settings is feasible and correlates well with other pandemic measures. Environmental surveillance may also detect high-risk areas in a setting and allow for estimation of the risk of transmission with high spatial resolution.

High touch surface monitoring has several advantages, making it an attractive monitoring strategy. This approach avoids bias introduced by variations in disease severity and test-seeking behavior. Additionally, task-shifting environmental surface sampling to individuals with minimal training makes this approach more accessible and scalable than individual testing by specialized clinical personnel. Since extensive human testing is impractical, high-touch surface monitoring may be beneficial at more minor spatial scales, such as within buildings. Surface sampling within a community might provide information on the locations of people who are now contagious and help with the early detection of SARS-CoV-2 instances when people are most contagious, such as when they are asymptomatic or pre-symptomatic (Arons et al., 2020; Lee et al., 2020). This may be especially helpful at primary schools and universities, where surfaces in each classroom and door entrances could be checked, and essential steps could be taken to decrease the possibility of spread. Surface surveillance has an added benefit of allowing analysis of swab samples using the same procedures and safety precautions as those used for human testing. Since cost-effective and faster human diagnostic tests are being developed during the pandemic, surface sampling could also take advantage of these advancements (Abdalhamid et al., 2020; Ben-Ami et al., 2020; Broughton et al., 2020; Lohse et al., 2020). In addition, this approach could be rapidly expanded to a variety of other pathogens.

Several factors were considered when choosing the sampling locations for our study, including the accessibility of the sampling sites throughout the year, the absence of automated doors, and areas with high numbers of students, employees, and visitors for classes, events, and sports. High traffic areas were intentionally chosen to improve our ability to detect SARS-CoV-2. It is worth noting that certain weeks were not sampled due to holidays, semester breaks, and hurricanes. Similar to other studies focused on environmental surveillance of SARS-CoV-2 (Hinz et al., 2022; Zuniga-Montanez et al., 2022), our research employed a weekly sampling schedule for systematic and representative collection of environmental samples over time. A study conducted by Ramuta et al. demonstrated that conducting weekly tests on environmental samples can yield comparable outcomes to daily sampling, reducing in the necessary testing resources, alleviating the overall workload, and offering valuable insights into the prevalence of pathogens during a specific period (Ramuta et al., 2022).

We employed qRT-PCR to identify SARS-CoV-2 RNA on high-touch environmental surface samples which provide semi-quantitative information on the viral gene copy present in the environmental surface. This method has been used in other studies to quantify viral gene copy on environmental surface (Mihajlovski et al., 2022; Zhang et al., 2022). However, these data should be interpreted carefully when collecting environmental surface samples from a community setting (Han et al., 2021). The viral RNA load on the environmental surfaces can be influenced by different variables, such as the location, the area swabbed, the usage of disinfectants on the surfaces, the quantity of virus shed by each infected person, the number of infected individuals, and the extent of the interior environment. With the size and complexity of a university, using an environmental surface swab may not adequately reflect the amount of viral gene copy in the environment. Surface sampling from numerous locations within a building may be necessary to detect the presence of infected persons and identify SARS-CoV-2 transmission hotspots more reliably. Further research is needed to determine how efficient this method is in various congregate settings.

The detection of viral RNA by RT-qPCR analysis cannot determine whether a contaminated surface contains infectious virus particle (Harvey et al., 2020). This may cause biases in settings where frequent disinfectant is used. Surface disinfectants can inactivate SARS-CoV-2 without largely impacting the detection of genetic material by conventional PCR (Krasnikova et al., 2022). Surface samples collected in these environments following disinfectant use could be positive by PCR without infectious viruses. Nonetheless, a nucleic acid-positive surface result in these settings could indicate that an infected individual was present in that place and might still pose a risk for transmission. We did not attempt to culture SARS-CoV-2 from the environmental surface in this study to determine if the infectious virus can be isolated from the nucleic acid-positive samples. Several studies have attempted to culture SARS-CoV-2 from the environmental surface, but most studies could not find viable or infectious viral in cell culture (Coil et al., 2021; Shragai et al., 2022).

During the study period, there was a significant rise in daily cases associated with the Omicron lineage wave in Florida. Throughout this period, university classrooms, laboratories, gymnasiums, student unions, and libraries were frequently used by students. We found that around 90% of environmental surfaces were positive for SARS-CoV-2 RNA, suggesting that indirect transmission through cross-contamination between students’ shared surfaces may have occurred in these spaces. The high positivity rate may be attributed to the sampling sites, which experienced frequent contact on a daily basis. This result differs from previous reports that analyzed environmental surfaces with lower positive rates (4.26–40%) (Harvey et al., 2020; Abrahão et al., 2021; Brazell et al., 2021; Casabianca et al., 2022). However, a direct comparison between our findings and those from similar studies is difficult since our study was conducted in areas with high individual touches. We found viral gene copy on door handles ranging from 0.4 to 60.6 gc/cm^2^, similar to other studies. Previously reported concentrations of SARS-CoV-2 RNA detected ranged from less than 0.1 to 40 gc/cm^2^ on public surfaces (Abrahão et al., 2021), 1.75 to 16.1 gc/cm^2^ on public transit surfaces (Brazell et al., 2021) and 0.84 gc/cm2 on bus terminal handrails (Moreno et al., 2021). Our findings contribute to a growing literature of detectable but low-level SARS-CoV-2 RNA contamination on public surfaces.

The comparative analysis of SARS-CoV-2 sequencing data between environmental swabs and clinical samples from university and state-level genomic surveillance revealed a significant overlap in the identified variants. This finding evidences the feasibility of generating sequencing data from environmental surface samples and suggests that employing this approach can determine the composition of circulating viral lineages. Of note, we documented instances where unique variants were identified in the environmental samples, which were also found in state clinical data but not in the clinical samples from student health center. This highlights the potential environmental sequencing to monitor circulating and novel variants with higher resolution. Our study also indicates the early detection of emerging variant is feasible by environmental sample sequencing. Early detection of Omicron variant through wastewater sequencing prior to its detection in clinical sequencing has been reported in multiple studies (Wilhelm et al., 2022a, 2022b; Gupta et al., 2023). In our study, BA. 5 was detected earlier in environmental samples than it was in university and state clinical data; however, we could not accurately estimate the lag time due to lack of consistent weekly environmental sequencing data. This high-resolution approach may be especially useful in locations with high host mobility (public transit, airports or attractions), where emerging variants may be detected, as well as settings with vulnerable populations (schools, nursing homes and hospitals), where spatially resolved monitoring is crucial for guiding interventions. Unlike clinical case samples, viral genome sequencing from ambient surfaces remains a technical difficulty due to low target molecule concentrations and nucleic acid degradation on dry surfaces (Nicholls et al., 2023). However, customized extraction method and sequencing techniques (Coil et al., 2021) are expected to provide higher resolution for detection and quantification of SARS-CoV-2 variants and other viruses.

This study is not without limitations. We did not have information on frequency and timing of surface disinfection, and the availability of hand sanitizer in the study area likely impacted the presence of SARS-CoV-2 RNA on surfaces and the strength of the association with COVID-19 cases. Multiple factors, including the number of students present on the campus, games, other events, holidays, and orientation programs, could impact the results of our study. We did not attempt to culture the virus from our surface samples and, therefore, cannot determine the viability or infectivity of the SARS-CoV-2 detected in our samples. Future work is needed to confirm the relationship between SARS-CoV-2 RNA concentrations and viable viruses on surfaces and to determine if infective SARS-CoV-2 can be recovered from environmental surface areas in community settings. Our study was conducted on a university campus, so extrapolating these findings to the general population or other non-healthcare settings should be done cautiously. Despite these limitations, our results are a valuable addition to the environmental surveillance with extension to a variety of other settings.

## Conclusion

This study investigated the extent of SARS-CoV-2 contamination on high touch surface areas in a university setting. We found that the gene copy number of SARS-CoV-2 in the environmental surfaces strongly correlated with the COVID-19 confirmed cases in the university. We also demonstrate the feasibility of identifying circulating or newly emerging SARS-CoV-2 variants in a community through viral genome sequencing from environmental surface samples. This non-invasive and cost-effective approach could be employed as an effective tool for monitoring SARS-CoV-2 and other infectious diseases.

## Data Availability

All data produced in the present study are available upon reasonable request to the authors

## Acknowledgement

We thank the global laboratories that generated and made public the SARS-CoV-2 sequences (through GISAID) used as reference dataset in this study (Supplementary Table S1).

## Ethics statement

This study was reviewed by the University of Central Florida Institutional Review Board and received a non-human subject determination.

## References

Abdalhamid B, Bilder CR, McCutchen EL, Hinrichs SH, Koepsell SA, Iwen PC (2020) Assessment of specimen pooling to conserve SARS CoV-2 testing resources. Am J Clin Pathol 153:715–718.

Abrahão JS, Sacchetto L, Rezende IM, Rodrigues RAL, Crispim APC, Moura C, Mendonça DC, Reis E, Souza F, Oliveira GFG (2021) Detection of SARS-CoV-2 RNA on public surfaces in a densely populated urban area of Brazil: A potential tool for monitoring the circulation of infected patients. Sci Total Environ 766:142645.

Ahmed W, Angel N, Edson J, Bibby K, Bivins A, O’Brien JW, Choi PM, Kitajima M, Simpson SL, Li J (2020) First confirmed detection of SARS-CoV-2 in untreated wastewater in Australia: a proof of concept for the wastewater surveillance of COVID-19 in the community. Sci Total Environ 728:138764.

Al Huraimel K, Alhosani M, Kunhabdulla S, Stietiya MH (2020) SARS-CoV-2 in the environment: Modes of transmission, early detection and potential role of pollutions. Sci Total Environ 744:140946.

Anon (n.d.) COVID-19: Wisconsin Coronavirus Wastewater Monitoring Network. Available at: https://dhs.wisconsin.gov/covid-19/wastewater.htm.

Arons MM, Hatfield KM, Reddy SC, Kimball A, James A, Jacobs JR, Taylor J, Spicer K, Bardossy AC, Oakley LP (2020) Presymptomatic SARS-CoV-2 infections and transmission in a skilled nursing facility. N Engl J Med 382:2081–2090.

Batista C, Hotez P, Amor Y Ben, Kim JH, Kaslow D, Lall B, Ergonul O, Figueroa JP, Gursel M, Hassanain M (2022) The silent and dangerous inequity around access to COVID-19 testing: a call to action. EClinicalMedicine 43.

Ben-Ami R, Klochendler A, Seidel M, Sido T, Gurel-Gurevich O, Yassour M, Meshorer E, Benedek G, Fogel I, Oiknine-Djian E (2020) Large-scale implementation of pooled RNA extraction and RT-PCR for SARS-CoV-2 detection. Clin Microbiol Infect 26:1248–1253.

Bowes DA, Driver EM, Kraberger S, Fontenele RS, Holland LA, Wright J, Johnston B, Savic S, Newell ME, Adhikari S (2023) Leveraging an established neighbourhood-level, open access wastewater monitoring network to address public health priorities: a population-based study. The Lancet Microbe 4:e29–e37.

Brazell LR, Stetz S, Hipp A, Taylor S, Stark N, Jensen K, Islam Juel MA, Deegan P, Munir M, Schlueter J (2021) Environmental screening for surface SARS-CoV-2 contamination in urban high-touch areas. medRxiv:2005–2021.

Brito AF, Semenova E, Dudas G, Hassler GW, Kalinich CC, Kraemer MUG, Ho J, Tegally H, Githinji G, Agoti CN (2022) Global disparities in SARS-CoV-2 genomic surveillance. Nat Commun 13:7003.

Broughton JP, Deng X, Yu G, Fasching CL, Servellita V, Singh J, Miao X, Streithorst JA, Granados A, Sotomayor-Gonzalez A (2020) CRISPR–Cas12-based detection of SARS-CoV-2. Nat Biotechnol 38:870–874.

Casabianca A, Orlandi C, Amagliani G, Magnani M, Brandi G, Schiavano GF (2022) SARS-CoV-2 RNA detection on environmental surfaces in a university setting of Central Italy. Int J Environ Res Public Health 19:5560.

Chen Z, Azman AS, Chen X, Zou J, Tian Y, Sun R, Xu X, Wu Y, Lu W, Ge S (2022) Global landscape of SARS-CoV-2 genomic surveillance and data sharing. Nat Genet:1–9.

Coil DA, Albertson T, Banerjee S, Brennan G, Campbell AJ, Cohen SH, Dandekar S, Díaz-Muñoz SL, Eisen JA, Goldstein T (2021) SARS-CoV-2 detection and genomic sequencing from hospital surface samples collected at UC Davis. PLoS One 16:e0253578.

Davies NG et al. (2021) Estimated transmissibility and impact of SARS-CoV-2 lineage B.1.1.7 in England. Science (80-) 372:eabg3055 Available at: http://science.sciencemag.org/content/372/6538/eabg3055.abstract.

Davó L, Seguí R, Botija P, Beltrán MJ, Albert E, Torres I, López-Fernández PÁ, Ortí R, Maestre JF, Sánchez G (2021) Early detection of SARS-CoV-2 infection cases or outbreaks at nursing homes by targeted wastewater tracking. Clin Microbiol Infect 27:1061–1063.

Duma Z, Chuturgoon AA, Ramsuran V, Edward V, Naidoo P, Mpaka-Mbatha MN, Bhengu KN, Nembe N, Pillay R, Singh R (2022) The challenges of severe acute respiratory syndrome coronavirus 2 (SARS-CoV-2) testing in low-middle income countries and possible cost-effective measures in resource-limited settings. Global Health 18:5.

for Immunization NC (2021) Science Brief: SARS-CoV-2 and Surface (Fomite) Transmission for Indoor Community Environments. In: CDC COVID-19 Science Briefs [Internet]. Centers for Disease Control and Prevention (US).

Gupta P, Liao S, Ezekiel M, Novak N, Rossi A, LaCross N, Oakeson K, Rohrwasser A (2023) Wastewater genomic surveillance captures early detection of Omicron in Utah. Microbiol Spectr:e00391–23.

Han MS, Byun J-H, Cho Y, Rim JH (2021) RT-PCR for SARS-CoV-2: quantitative versus qualitative.

Harvey AP, Fuhrmeister ER, Cantrell ME, Pitol AK, Swarthout JM, Powers JE, Nadimpalli ML, Julian TR, Pickering AJ (2020) Longitudinal monitoring of SARS-CoV-2 RNA on high-touch surfaces in a community setting. Environ Sci Technol Lett 8:168–175.

Hassouneh SA-D, Trujillo A, Ali S, Cella E, Johnston C, DeRuff KC, Sabeti PC, Azarian T (2023) Antigen test swabs are comparable to nasopharyngeal swabs for sequencing of SARS-CoV-2. Sci Rep 13:11255 Available at: https://doi.org/10.1038/s41598-023-37893-5.

Heneghan CJ, Spencer EA, Brassey J, Plüddemann A, Onakpoya IJ, Evans DH, Conly JM, Jefferson T (2021) SARS-CoV-2 and the role of orofecal transmission: A systematic review. F1000Research 10.

Hinz A, Xing L, Doukhanine E, Hug LA, Kassen R, Ormeci B, Kibbee RJ, Wong A, MacFadden D, Nott C (2022) SARS-CoV-2 detection from the built environment and wastewater and its use for hospital surveillance. Facets 7:82–97.

Jahn K, Dreifuss D, Topolsky I, Kull A, Ganesanandamoorthy P, Fernandez-Cassi X, Bänziger C, Devaux AJ, Stachler E, Caduff L (2022) Early detection and surveillance of SARS-CoV-2 genomic variants in wastewater using COJAC. Nat Microbiol 7:1151–1160.

Karthikeyan S, Levy JI, De Hoff P, Humphrey G, Birmingham A, Jepsen K, Farmer S, Tubb HM, Valles T, Tribelhorn CE (2022) Wastewater sequencing reveals early cryptic SARS-CoV-2 variant transmission. Nature 609:101–108.

Krasnikova MS, Lazareva EA, Yatsentyuk SP (2022) The Effect of Disinfectants on the SARS-CoV-2 RNA Detection in Swabs from Various Surfaces. Appl Biochem Microbiol 58:932–937.

Kwon T, Osterrieder N, Gaudreault NN, Richt JA (2023) Fomite Transmission of SARS-CoV-2 and Its Contributing Factors. Pathogens 12:364.

Lee S, Kim T, Lee E, Lee C, Kim H, Rhee H, Park SY, Son H-J, Yu S, Park JW (2020) Clinical course and molecular viral shedding among asymptomatic and symptomatic patients with SARS-CoV-2 infection in a community treatment center in the Republic of Korea. JAMA Intern Med 180:1447–1452.

Leifels M, Lee WL, Armas F, Gu X, Chandra F, Cheng D, Kwok WC, Chua D, Kim SY, Ng WJ (2023) Surveillance of SARS-CoV-2 in Wastewater at the Population Level: Insights into the Implementation of Non-invasive Targeted Monitoring in Singapore and the USA. In. Springer.

Lieberman-Cribbin W, Tuminello S, Flores RM, Taioli E (2020) Disparities in COVID-19 testing and positivity in New York City. Am J Prev Med 59:326–332.

Lohse S, Pfuhl T, Berkó-Göttel B, Rissland J, Geißler T, Gärtner B, Becker SL, Schneitler S, Smola S (2020) Pooling of samples for testing for SARS-CoV-2 in asymptomatic people. Lancet Infect Dis 20:1231–1232.

Lu X, Wang L, Sakthivel SK, Whitaker B, Murray J, Kamili S, Lynch B, Malapati L, Burke SA, Harcourt J (2020) US CDC real-time reverse transcription PCR panel for detection of severe acute respiratory syndrome coronavirus 2. Emerg Infect Dis 26:1654.

Lucas C, Vogels CBF, Yildirim I, Rothman JE, Lu P, Monteiro V, Gehlhausen JR, Campbell M, Silva J, Tabachnikova A (2021) Impact of circulating SARS-CoV-2 variants on mRNA vaccine-induced immunity. Nature 600:523–529.

Mihajlovski K, Buttner MP, Cruz P, Labus B, St. Pierre Schneider B, Detrick E (2022) SARS-CoV-2 surveillance with environmental surface sampling in public areas. PLoS One 17:e0278061.

Moreno T, Pintó RM, Bosch A, Moreno N, Alastuey A, Minguillón MC, Anfruns-Estrada E, Guix S, Fuentes C, Buonanno G, Stabile L, Morawska L, Querol X (2021) Tracing surface and airborne SARS-CoV-2 RNA inside public buses and subway trains. Environ Int 147:106326 Available at: https://www.sciencedirect.com/science/article/pii/S0160412020322819.

Nicholls IG, Spencer A, Chen Y, Bennett A, Atkinson B (2023) Surface sampling for SARS-CoV-2 in workplace outbreak settings in the UK, 2021-22. medRxiv:2002–2023.

Posit Team (2023) RStudio: Integrated Development Environment for R. Posit Software, PBC, Boston, MA.

Ramuta MD, Newman CM, Brakefield SF, Stauss MR, Wiseman RW, Kita-Yarbro A, O’Connor EJ, Dahal N, Lim A, Poulsen KP (2022) SARS-CoV-2 and other respiratory pathogens are detected in continuous air samples from congregate settings. Nat Commun 13:4717.

Reitsma MB, Claypool AL, Vargo J, Shete PB, McCorvie R, Wheeler WH, Rocha DA, Myers JF, Murray EL, Bregman B (2021) Racial/Ethnic Disparities In COVID-19 Exposure Risk, Testing, And Cases At The Subcounty Level In California: Study examines racial/ethnic disparities in COVID-19 risk, testing, and cases. Health Aff 40:870–878.

Shragai T, Pratt C, Castro Georgi J, Donnelly MAP, Schwartz NG, Soto R, Chuey M, Chu VT, Marcenac P, Park GW (2022) Household characteristics associated with surface contamination of SARS-CoV-2 and frequency of RT-PCR and viral culture positivity–California and Colorado, 2021. PLoS One 17:e0274946.

Smyth DS, Trujillo M, Gregory DA, Cheung K, Gao A, Graham M, Guan Y, Guldenpfennig C, Hoxie I, Kannoly S (2022) Tracking cryptic SARS-CoV-2 lineages detected in NYC wastewater. Nat Commun 13:635.

Wickham H, Chang W, Wickham MH (2016) Package ‘ggplot2.’ Creat elegant data Vis using Gramm Graph Version 2:1–189.

Wilhelm A, Agrawal S, Schoth J, Meinert-Berning C, Bastian D, Orschler L, Ciesek S, Teichgräber B, Wintgens T, Lackner S (2022a) Early detection of SARS-CoV-2 Omicron BA. 4 and BA. 5 in German wastewater. Viruses 14:1876.

Wilhelm A, Schoth J, Meinert-Berning C, Agrawal S, Bastian D, Orschler L, Ciesek S, Teichgräber B, Wintgens T, Lackner S (2022b) Wastewater surveillance allows early detection of SARS-CoV-2 omicron in North Rhine-Westphalia, Germany. Sci Total Environ 846:157375.

Zhang X, Wu J, Smith LM, Li X, Yancey O, Franzblau A, Dvonch JT, Xi C, Neitzel RL (2022) Monitoring SARS-CoV-2 in air and on surfaces and estimating infection risk in buildings and buses on a university campus. J Expo Sci Environ Epidemiol 32:751–758.

Zuniga-Montanez R, Coil DA, Eisen JA, Pechacek R, Guerrero RG, Kim M, Shapiro K, Bischel HN (2022) The challenge of SARS-CoV-2 environmental monitoring in schools using floors and portable HEPA filtration units: Fresh or relic RNA? PLoS One 17:e0267212.

